# Relationship of TSH levels with cardiometabolic risk factors in US youth aged 12-18 years and population-based reference percentiles for thyroid function tests

**DOI:** 10.1101/2020.09.20.20198341

**Authors:** Xinlei Chen, Shuliang Deng, Cecilia Sena, Chuhan Zhou, Vidhu V. Thaker

## Abstract

**Context:** Thyroid hormones play an important role in the metabolic homeostasis of the body and have been associated with cardiometabolic risk.

**Objective:** To examine the association of cardiometabolic risk factors (CMRF) with TSH levels in youth at population level in the US.

**Design & Setting:** Cross-sectional study of youth aged 12-18 years without known thyroid abnormalities from National Health and Nutrition Examination Survey 1999-2012. Subclinical hypothyroidism (SH) was defined as TSH levels 4.5-10 mIU/L. Assessed CMRF included abdominal obesity (waist circumference > 90^th^percentile), hypertriglyceridemia (TG ≥ 130 mg/dL), low HDL cholesterol (HDL-C < 40 mg/dL), elevated blood pressure (SBP and DBP ≥ 90^th^percentile), hyperglycemia (FBG ≥ 100 mg/dL, or known diabetes), insulin resistance (HOMA-IR > 3.16) and elevated alanine transferase (ALT ≥50 U/L for boys and ≥44 U/L for girls). Age and sex-specific percentiles for thyroid parameters were calculated for youth with normal weight.

**Results:** In this cohort of youth (51.3% male), 31.2% had overweight/obesity. The prevalence of SH was 2.0 % (95% CI 1.2-3.1). The median TSH levels were higher in youth with overweight/obesity (p<.001). Adjusting for age, sex, race/ethnicity and level of obesity, youth with TSH in the 4^th^ quantile had higher odds of abdominal obesity (OR 2.53 [1.43-4.46], p = .002), higher HOMA-IR (OR 2.82 [1.42-5.57], p=.003) and ≥ 2 CMRF (OR 2.20 [1.23-3.95], p=.009).

**Conclusions:** The prevalence of SH is low in US youth. The higher odds of insulin resistance and CMRF in youth with TSH levels > 75^th^ percentile requires further study.

## Introduction

Thyroid hormones play an important role in normal growth and development as well as regulating metabolism, thermogenesis and cardiovascular function (1). Many of the metabolic activities regulated by thyroid hormones are related to the anabolism and/or catabolism of macromolecules that affect energy homeostasis, and thyroid hormones triiodothyronine (T3) and thyroxine (T4) have direct effects on both cholesterol and fatty acid synthesis and metabolism (2,3). The feedback loop of thyroid function is regulated by the thyroid stimulating hormone (TSH) secreted from the anterior pituitary in response to the circulating thyroid hormone(s). TSH levels in the blood are the most sensitive markers of optimal thyroid function and mild elevation of TSH, called subclinical hypothyroidism (SH, TSH 4.5-10 mIU/L), has been considered the earliest manifestation of thyroid dysfunction. In adults, SH has been associated with adverse cardiovascular outcomes, such as carotid intima thickness (4,5), coronary heart disease (6), heart failure (7), stroke (8), non-alcoholic fatty liver disease (9), and the role of thyroid replacement therapy is being debated (10,11).

Cohort studies in children and adolescents have found an association between elevated TSH levels or SH and obesity (12-17). An association has also been demonstrated between SH and cardiometabolic risk factors (CMRF), such as hypercholesterolemia (18,19), carotid intima thickness (19) and glucose dysmetabolism (20-22). Whether these associations are a result of the insulin resistance seen with obesity or are independently associated with TSH remains to be established. There is sparsity of such studies from population-based cohorts.

The objective of this study was to assess the relationship between TSH levels and CMRF namely abdominal obesity, dyslipidemia, glucose intolerance, elevated blood pressure, insulin resistance and elevated alanine transferase (23) in a nationally representative sample of children and adolescents from the US from the National Health and Nutrition Examination Survey (NHANES). Additionally, we aimed to identify the reference percentiles for measures of thyroid function by age and gender in children with BMI percentiles between 5-85.

## Material and Methods

### Study population

The data for this study was obtained from combining five 2-year cycles of NHANES where thyroid function tests were measured (1999-2000; 2001-2002 and 2007-2008 through 2011-2012). NHANES is a cross-sectional, multi-stage complex nationally representative survey that uses interview, physical examination and laboratory data to assess the health and nutritional status of the non-institutionalized civilian population in the US(24).

### Study cohort

Subjects for the study are youth between the ages of 12-18 years at the time of examination. Data pertinent to the thyroid function, lipid levels, liver enzymes, measures of glucose homeostasis in fasting samples, anthropometric and demographic measures were obtained. Subjects with history of thyroid disease or those on thyroid supplements or drugs that influence thyroid function, such as steroids, lithium, amiodarone, or beta blockers were excluded. Additionally, subjects with clinical hypothyroidism, hyperthyroidism, the presence of anti-thyroid antibodies (anti-thyroidperoxidase [TPO] or anti-thyroglobulin [TG]) and those with abnormal creatinine were excluded from the analysis. The interview data contains information on race/ethnicity that was classified as non-Hispanic white (NHW), non-Hispanic black (NHB), Hispanic or other. The average response rate for the study cohort was 80% (range 75 – 85). The National Center for Health Statistics Institutional Review Board oversaw the written, informed consent for emancipated minors and parents and guardians for minors for data collection.

### Anthropometric measurement

NHANES survey instruments and protocols have been established and ongoing in 2-year cycles since 1999 (25). Briefly, anthropometric measures include weight measured to the nearest 0.1kg using a digital weight scale with participants wearing the standard mobile examination center (MEC) examination gown. Standing height was measured to the nearest 0.1cm using a stadiometer with a fixed vertical backboard and an adjustable head piece. Body mass index (BMI) was calculated as weight (kg)/height (m) squared. Subjects with BMI between 5-84 percentile were considered without obesity, 85-94 percentile overweight and ≥ 95^th^ percentile with obesity (26). Waist circumference was measured in the horizontal plane to the nearest 0.1cm at the junction between the uppermost lateral border of the right ilium and the midaxillary line which extends from the armpit down the side of the torso. Blood pressure was measured 3 times and the average was used.

### Biochemistry assessment

TSH was measured with a two-site immunoenzymatic assay and the total T4 was measured with a competitive binding immunoenzymatic assay (27-31) (eTable 1). The Total T4 levels for 2011-2012 cycle were adjusted to align with the measurements from the prior years for instrument change (31). TG antibody and TPO antibody were measured using a sequential two-step immunoenzymatic “sandwich” assay (28-31). Triglycerides were measured enzymatically in serum or plasma using a series of coupled reactions in which triglycerides are hydrolyzed to produce glycerol. High density cholesterol (HDL-C) was measured by heparin-manganese precipitation or direct immunoassay method. The fasting plasma glucose and insulin levels were forward calibrated to the methodology used in 2011-12 (32-36). Alanine Aminotransferase (ALT) was measured by the catalytic action that causes the interconversion of amino acids and *α*-ketoacids by transfer of amino groups.

### Cardiometabolic risk factors (CMRF)

We used the organizational framework offered by metabolic syndrome (MetS) to assess the CMRF in children and adolescents (23). For this study, the criteria defined by the International Diabetes Federation (37) were updated for hypertriglyceridemia and elevated blood pressure according to current guidelines as follows: a) abdominal obesity (waist circumference > 90th percentile for 10-16 years and/or ≥ 94 cm in males and ≥ 80 cm for females); b) hypertriglyceridemia (TG ≥ 130 mg/dL) (38); c) low high density cholesterol (HDL-C < 40 mg/dL); d) elevated blood pressure (Systolic and Diastolic BP ≥ 90^th^ percentile) (39); d) hyperglycemia (FBG ≥ 100 mg/dL, or reported diagnosis of diabetes). Two additional measures of metabolic risk used were insulin resistance measured by homeostatic model for insulin resistance (HOMA-IR = fasting insulin (microU/L) x fasting glucose (mmol/L) /22.5) (40) with > 3.16 considered high (41) and ALT levels as a measure for hepatic steatosis (≥50 U/L for boys and ≥44 U/L for girls) (42).

### Data analysis

BMI and waist circumference were transformed into age and sex appropriate percentiles and z-score respectively using the CDC 2000 growth charts (43). The thyroid related variables were examined by probability density estimation to assess the shape of distribution. The summary of the distribution is presented as median with standard deviation for continuous variables or percentage for categorical variables. The characteristics of participants were compared using complex survey design adjusted differences in the median.

TSH quantiles were generated using quantile regression adjusting for age and sex to account for the absence of the pubertal status in the NHANES dataset. The 4^th^ quantile was considered to be relevant for cardiometabolic risk. After ensuring absence of temporal trends between the survey cycles, the 5-survey cycles were combined for analysis and 10-year survey weights were constructed by rescaling the NHANES survey weights so that the sum of the weights matches the survey population at the midpoint of that period(44). Univariate and multivariable analysis using complex survey-based generalized linear models with a logarithmic link was used to assess the odds of the CMRF with TSH quantiles while adjusting for the age, gender, race/ethnicity and level of obesity. A binary classification of TSH using the definition of SH (4.5 mIU/L with normal free T4 levels) was used to assess the association of the risks with SH (10,45). Survey based goodness of fit tests were done to test the model fit. Interaction between the predictors were tested using design-adjusted Wald test and likelihood ratio tests.

We imputed the missing data of TSH by multivariate imputation by chained equations using predictive mean matching to ascertain the percentiles of TSH. The imputation was verified by random verification of the complete dataset with similar methods. Sensitivity analysis for the results was performed using the complete cases. The terms with p-value < 0.05 were considered statistically significant and 95% confidence interval (CI) was calculated. Age- and sex-specific percentiles of TSH levels and other thyroid parameters among subjects with BMI between 5-85^th^ percentiles were calculated using the imputed dataset by Generalized Additive Models for Location Scale and Shape (GAMLSS) package using different Box-Cox transformation distribution family and the best fitted model was determined by the Akaike Information Criterion.

Statistical analyses were performed in R v3.5.1 using tidyverse, survey, quantreg, mice, gamlss packages and STATA (StataCorp 2019. *Stata Statistical Software: Release 16*. College Station, TX: StataCorp LLC).

## Results

### Characteristics of study subjects

The final study cohort was representative of 15.4 million youth between the ages of 12-18 years of age (51.3% boys, 95% CI 49.2-53.4%), of which 31.2% (95% CI 28.5-38.1%) had BMI ≥ 85^th^ percentile. Participants were excluded if TSH ≥ 10 mIU/L (n = 5), total T4 > 13.2 ug/dL (n= 10), positive for antithyroid antibodies (n = 202), or on medications affecting thyroid function (n = 20). The characteristics of the study subjects are shown in Table 1 grouped by BMI percentile. The distribution of age and gender was similar between the two groups. As has been shown before, the prevalence of obesity was higher in Hispanic and Non-Hispanic Black youth(46). The CMRF were higher in the group with overweight/obesity by BMI percentile. The TSH levels were significantly different between the two groups, while the total and free T4 and total T3 levels were not. In the subgroup analysis by gender, the TSH levels were similar in both, but other thyroid parameters were statistically different. In the CMRF, waist circumference z-scores were lower in girls with obesity. HDL-C levels were higher and ALT levels were lower in the girls, and HOMA-IR was similar in either gender with obesity (eTable 2). We evaluated the temporal trends in TSH, Total T4 and Total T3 by NHANES cycles and found no differences (eFigure 1). The distribution of the CMRF by the TSH quantiles and gender is shown in eTable 3.

**Table 1.** Demographic distribution of the cohort.

|  | **BMI < 85 %tile**<br>**Median (95% CI)** | **BMI ≥ 85 %tile**<br>**Estimate (95% CI)** | **p-value** |
| --- | --- | --- | --- |
| **Proportion of population, %** | 68.7 (65.9-71.5) | 31.2 (28.5-34.1) |  |
| **Male, %** | 51.3 (48.8-53.8) | 52.2 (48.1- 56.2) | .73 |
| **Age, mean (years)** | 15.3 (15.2-15.5) | 15.2 (14.9-15.4) | .28 |
| **Race/ethnicity, %** |  |  |  |
| Hispanic | 15.7 (12.8-19.3) | 22.9 (18.1-28.6) | .001 |
| Non-Hispanic Black | 12.7 (10.6-15.1) | 20.2 (16.3-24.7) | <.001 |
| Non-Hispanic White | 63.0 (59.1-66.8) | 50.5 (44.3-56.7) | <.001 |
| **BMI z-score** | 0.07 (0.02-0.13) | 1.76 (1.71-1.80) | <.001 |
| **Waist circumference z-score** | -0.05 (-0.02-0.08) | 1.42 (1.37-1.46) | <.001 |
| **Abdominal obesity, %** | 6.6 (5.1-8.7) | 92.9 (90.8-94.5) | <.001 |
| **Waist-height ratio > 0.5, %** | 12.3 (9.2-16.3) | 93.0 (90.8-94.7) | <.001 |
| **Systolic BP, mm Hg** | 107.0 (106.5-107.5) | 112.0 (111.0-113.0) | <.001 |
| **TSH (uIU/mL)** | 1.33 (1.28-1.39) | 1.49 (1.43-1.54) | <.001 |
| **Total T4 (ug/dL)** | 7.50 (7.45-7.55) | 7.60 (7.50-7.70) | .07 |
| **Total T3 (ng/dL)** | 129.0 (127.0-131.0) | 131.5 (129.0-133.9) | .12 |
| **Free T4 (ng/dL)** | 0.80 (0.78-0.81) | 0.80 (0.77-0.81) | .31 |
| **Free T3 (pg/mL)** | 3.63 (3.60-3.65) | 3.70 (3.65-3.75) | .009 |
| **Triglyceride (mg/dL)** | 66.0 (63.1-68.9) | 83.0 (77.6-88.4) | <.001 |
| **HDL-cholesterol (mg/dL)** | 52.0 (51.0-53.0) | 45.0 (44.0-46.0) | <.001 |
| **HOMA-IR** | 1.74 (1.66-1.81) | 3.24 (3.01-3.46) | <.001 |
| **ALT (U/L)** | 15.0 (14.5-15.5) | 19.0 (18.5-19.5) | <.001 |

Abbreviations: BMI = body mass index; BP= blood pressure; TSH= thyroid stimulating hormone; T4 = thyrotropin; T3= thyroxine; HDL = high density lipoprotein; HOMA-IR = homeostatic model for insulin resistance, ALT = alanine transferase
SI unit conversion: Total T4 (nmol/L) = multiply by 12.9; Total T3 (nmol/L) = multiply by 0.0154; fT4 (pmol/L) = multiply by 12.9;
fT3 (pmol/L) = multiply by 1.5362; TG (mmol/L) = multiply by 0.0113; HDL-C (mmol/L) = multiply by 0.0259
<sup>a</sup>BMI percentile adjusted for age and sex

### Cardiometabolic risk factors by TSH levels

The CMRF were significantly higher in the group with overweight/obesity by BMI percentile, as were the TSH levels (Table 1). There was a positive correlation between TSH levels and BMI-z score (r = 0.1, p <.001), waist circumference (r=0.2, p<.001), HOMA-IR (r= 0.1, p <.001), and triglycerides (r=0.1, p <.001). A greater number of subjects with overweight and obesity were seen in the higher age and sex adjusted quantiles (Fig. 1). In the regression analyses weighted by the 10-year constructed weights, the 4^th^ quantile of TSH was associated with increased odds of abdominal obesity, hypertriglyceridemia, elevated blood pressure, abnormal HOMA-IR, abnormal ALT levels and multiple risk factors as compared to the 1^st^ quantile (Table 2, Model 1). This association continued to have statistical significance while controlling for age, gender and race/ethnicity (Model 2). When controlling for level of obesity in addition to age, gender and race/ethnicity, the 4^th^ quantile for TSH was significantly associated with abdominal obesity, abnormal HOMA-IR and ≥ 2 CMRF (Model 3). Two percent (95% CI 1.2-3.1) subjects had SH by the conventional definition, with no differences between the two BMI categories. No association was seen between the presence of SH and any of the CMRF. No statistically significant interactions were identified between the predictors using design-based tests.

**Table 2.** Association between 4^th^ quantile of TSH and cardiometabolic risk factors.

| Regression models | Subpopulation size<br>(population represented) | Model 1 |  | Model 2 |  | Model 3 |  |
| --- | --- | --- | --- | --- | --- | --- | --- |
|  |  | OR*<br>(95% CI) | p-value | OR*<br>(95% CI) | p-value | OR*<br>(95% CI) | p-value |
| Abdominal obesity | 2509<br>(13.6 million) | 3.47<br>(2.37-5.10) | **<.001** | 3.57<br>(2.42-5.25) | **<.001** | 2.53<br>(1.43-4.46) | **.002** |
| Hypertriglyceridemia | 1118<br>(12.1 million) | 2.08<br>(1.01-4.27) | **.04** | 2.12<br>(1.00-4.92) | .05 | 1.84<br>(0.84-4.00) | .12 |
| Low HDL cholesterol | 2548<br>(13.8 million) | 1.03<br>(0.67-1.58) | .89 | 1.02<br>(0.66-1.58) | .92 | 0.81<br>(0.51-1.29) | .37 |
| Elevated blood pressure | 2199<br>(11.9 million) | 1.74<br>(1.05-2.87) | **.03** | 1.98<br>(1.21-3.23) | **.007** | 1.65<br>(1.00-2.73) | .05 |
| Hyperglycemia | 1118<br>(12.1 million) | 1.33<br>(0.73-2.41) | .35 | 1.15<br>(0.64-2.07) | .63 | 1.15<br>(0.62-2.11) | .65 |
| HOMA-IR | 1112<br>(12.0 million) | 3.34<br>(1.91-5.85) | **<.001** | 3.50<br>(1.98-6.17) | **<.001** | 2.82<br>(1.42-5.57) | **.003** |
| Elevated ALT level | 2546<br>(13.9 million) | 1.65<br>(0.48-5.66) | .42 | 1.79<br>(0.54-5.95) | .97 | 1.02<br>(0.33-3.17) | .97 |
| Risk score | 1118<br>(12.1 million) | 2.41<br>(1.42-4.07) | **.001** | 2.30<br>(1.34-3.97) | **.003** | 2.20<br>(1.23-3.95) | **.009** |

Abbreviations: OR = odds ratio, HDL = high-density lipoprotein, HOMA-IR = homeostatic model for insulin resistance, ALT = alanine transferase TSH quantile defined by quantile regression model adjusted for age and sex. 1<sup>st</sup> Quantile used as reference.
Model 1= unadjusted; Model 2= adjusted for age, sex and race/ethnicity; Model 3 = adjusted for age, sex, race/ethnicity and BMI

**Figure 1.**
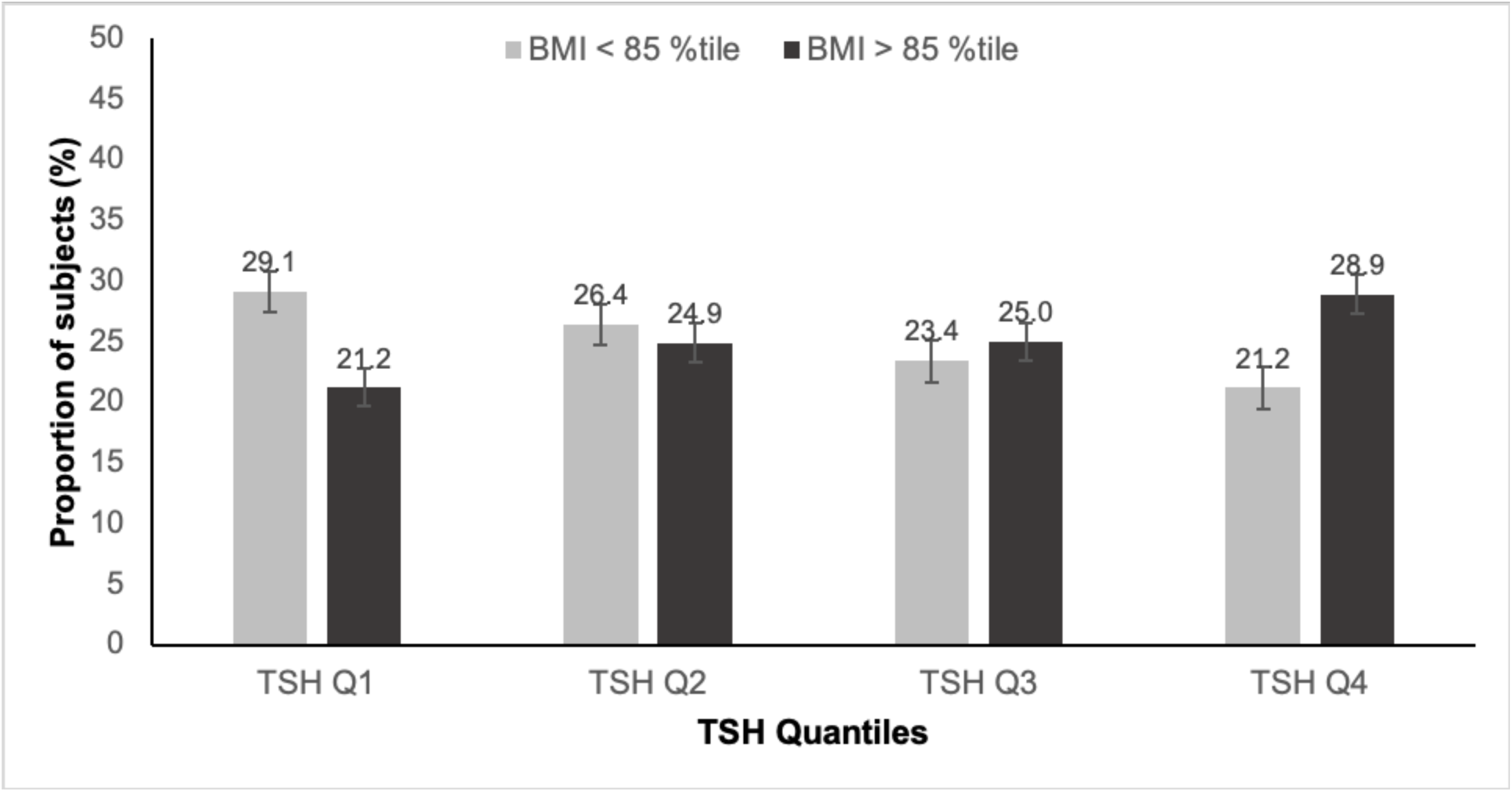
Proportion of subjects in each quantile of TSH based on the BMI percentile.

### Age- and sex-specific percentile of TSH levels among normal weight subjects

Generalized Additive Models were used to derive the age- and sex-specific percentiles of thyroid parameters based on the values for the subjects with BMI between 5-85^th^ percentile (Table 3). TSH levels decreased with increasing age and were consistently lower in girls compared to the boys in each age category. On the other hand, the total T4 levels increased with age and were higher in girls compared to the boys. The total T3 levels decreased with increasing age and were higher in girls. The levels of the free T4 as well as free T3 remained remarkably consistent across ages. The percentile curves can be seen in eFigure 2.

**Table 3.** Age- and sex-specific percentiles for thyroid parameters.

TSH level (μU/mL)
| Age<br>(years) | Percentiles for boys |  |  |  |  |  |  |  | Percentiles for girls |  |  |  |  |  |  |  |
| --- | --- | --- | --- | --- | --- | --- | --- | --- | --- | --- | --- | --- | --- | --- | --- | --- |
|  | Sample<br>size | 2.5 | 5 | 25 | 50 | 75 | 95 | 97.5 | Sample<br>size | 2.5 | 5 | 25 | 50 | 75 | 95 | 97.5 |
| 12 | 111 | 0.5 | 0.6 | 1.1 | 1.6 | 2.2 | 3.6 | 4.3 | 106 | 0.4 | 0.5 | 1.0 | 1.4 | 2.1 | 3.8 | 4.8 |
| 13 | 132 | 0.5 | 0.6 | 1.1 | 1.6 | 2.2 | 3.6 | 4.3 | 150 | 0.4 | 0.5 | 0.9 | 1.4 | 2.0 | 3.7 | 4.6 |
| 14 | 135 | 0.5 | 0.6 | 1.1 | 1.5 | 2.2 | 3.6 | 4.2 | 158 | 0.4 | 0.5 | 0.9 | 1.3 | 1.9 | 3.5 | 4.4 |
| 15 | 142 | 0.4 | 0.6 | 1.0 | 1.4 | 2.0 | 3.3 | 4.0 | 122 | 0.4 | 0.5 | 0.9 | 1.3 | 1.8 | 3.4 | 4.2 |
| 16 | 143 | 0.4 | 0.5 | 1.0 | 1.4 | 1.9 | 3.1 | 3.7 | 132 | 0.4 | 0.5 | 0.8 | 1.2 | 1.8 | 3.2 | 4.1 |
| 17 | 131 | 0.4 | 0.5 | 0.9 | 1.4 | 1.9 | 3.1 | 3.7 | 123 | 0.4 | 0.4 | 0.8 | 1.1 | 1.7 | 3.1 | 3.9 |
| 18 | 100 | 0.4 | 0.5 | 1.0 | 1.4 | 2.0 | 3.2 | 3.9 | 109 | 0.3 | 0.4 | 0.8 | 1.1 | 1.6 | 3.0 | 3.7 |

Total T4 level (μg/dL)
| Age<br>(years) | Percentiles for boys |  |  |  |  |  |  |  | Percentiles for girls |  |  |  |  |  |  |  |
| --- | --- | --- | --- | --- | --- | --- | --- | --- | --- | --- | --- | --- | --- | --- | --- | --- |
|  | Sample<br>size | 2.5 | 5 | 25 | 50 | 75 | 95 | 97.5 | Sample<br>size | 2.5 | 5 | 25 | 50 | 75 | 95 | 97.5 |
| 12 | 92 | 5.3 | 5.6 | 6.6 | 7.4 | 8.3 | 9.8 | 10.0 | 81 | 5.3 | 5.6 | 6.8 | 7.6 | 8.5 | 10.0 | 11.0 |
| 13 | 115 | 5.2 | 5.5 | 6.5 | 7.2 | 8.1 | 9.6 | 10.0 | 114 | 5.3 | 5.6 | 6.7 | 7.5 | 8.4 | 10.0 | 11.0 |
| 14 | 113 | 5.1 | 5.4 | 6.4 | 7.1 | 7.9 | 9.4 | 10.0 | 130 | 5.3 | 5.6 | 6.7 | 7.5 | 8.4 | 10.0 | 11.0 |
| 15 | 113 | 5.1 | 5.4 | 6.4 | 7.1 | 8.0 | 9.4 | 10.0 | 98 | 5.3 | 5.7 | 6.8 | 7.6 | 8.5 | 10.0 | 11.0 |
| 16 | 122 | 5.1 | 5.5 | 6.4 | 7.2 | 8.1 | 9.5 | 10.0 | 112 | 5.4 | 5.8 | 6.9 | 7.7 | 8.7 | 10.0 | 11.0 |
| 17 | 114 | 5.2 | 5.5 | 6.5 | 7.3 | 8.2 | 9.7 | 10.0 | 100 | 5.5 | 5.9 | 7.1 | 7.9 | 8.8 | 11.0 | 11.0 |
| 18 | 89 | 5.3 | 5.6 | 6.6 | 7.4 | 8.3 | 9.8 | 10.0 | 94 | 5.7 | 6.0 | 7.2 | 8.1 | 9.0 | 11.0 | 12.0 |

Total T3 level (ng/dL)
| Age<br>(years) | Percentiles for boys |  |  |  |  |  |  |  | Percentiles for girls |  |  |  |  |  |  |  |
| --- | --- | --- | --- | --- | --- | --- | --- | --- | --- | --- | --- | --- | --- | --- | --- | --- |
|  | Sample<br>size | 2.5 | 5 | 25 | 50 | 75 | 95 | 97.5 | Sample<br>size | 2.5 | 5 | 25 | 50 | 75 | 95 | 97.5 |
| 12 | 61 | 112 | 118 | 137 | 151 | 166 | 191 | 200 | 65 | 99 | 105 | 125 | 138 | 154 | 185 | 197 |
| 13 | 91 | 108 | 114 | 132 | 146 | 160 | 184 | 193 | 89 | 91 | 97 | 115 | 128 | 142 | 171 | 182 |
| 14 | 87 | 104 | 110 | 128 | 141 | 154 | 178 | 186 | 96 | 87 | 93 | 110 | 122 | 136 | 163 | 175 |
| 15 | 93 | 100 | 105 | 123 | 135 | 149 | 171 | 179 | 76 | 85 | 91 | 108 | 120 | 133 | 159 | 170 |
| 16 | 91 | 96 | 101 | 118 | 130 | 143 | 164 | 172 | 84 | 85 | 90 | 107 | 118 | 132 | 158 | 169 |
| 17 | 91 | 92 | 97 | 113 | 125 | 137 | 158 | 165 | 79 | 85 | 91 | 108 | 120 | 133 | 159 | 170 |
| 18 | 72 | 88 | 93 | 108 | 119 | 131 | 151 | 158 | 69 | 88 | 94 | 111 | 124 | 137 | 165 | 176 |

Free T4 level (ng/dL)
| Age<br>(years) | Percentiles for boys |  |  |  |  |  |  |  | Percentiles for girls |  |  |  |  |  |  |  |
| --- | --- | --- | --- | --- | --- | --- | --- | --- | --- | --- | --- | --- | --- | --- | --- | --- |
|  | Sample<br>size | 2.5 | 5 | 25 | 50 | 75 | 95 | 97.5 | Sample<br>size | 2.5 | 5 | 25 | 50 | 75 | 95 | 97.5 |
| 12 | 61 | 0.6 | 0.6 | 0.7 | 0.8 | 0.9 | 1.0 | 1.1 | 65 | 0.6 | 0.6 | 0.7 | 0.8 | 0.9 | 1.0 | 1.1 |
| 13 | 91 | 0.6 | 0.6 | 0.7 | 0.8 | 0.9 | 1.0 | 1.1 | 89 | 0.6 | 0.6 | 0.7 | 0.8 | 0.9 | 1.0 | 1.1 |
| 14 | 87 | 0.6 | 0.6 | 0.7 | 0.8 | 0.9 | 1.0 | 1.1 | 97 | 0.6 | 0.6 | 0.7 | 0.8 | 0.9 | 1.0 | 1.1 |
| 15 | 93 | 0.6 | 0.6 | 0.7 | 0.8 | 0.9 | 1.0 | 1.1 | 76 | 0.6 | 0.6 | 0.7 | 0.8 | 0.9 | 1.0 | 1.1 |
| 16 | 91 | 0.6 | 0.6 | 0.7 | 0.8 | 0.9 | 1.0 | 1.1 | 84 | 0.6 | 0.6 | 0.7 | 0.8 | 0.9 | 1.0 | 1.1 |
| 17 | 91 | 0.6 | 0.6 | 0.7 | 0.8 | 0.9 | 1.0 | 1.1 | 79 | 0.6 | 0.6 | 0.7 | 0.8 | 0.9 | 1.0 | 1.1 |
| 18 | 72 | 0.6 | 0.6 | 0.7 | 0.8 | 0.9 | 1.0 | 1.1 | 69 | 0.6 | 0.6 | 0.7 | 0.8 | 0.9 | 1.0 | 1.1 |

Free T3 level (pg/mL)
| Age<br>(years) | Percentiles for boys |  |  |  |  |  |  |  | Percentiles for girls |  |  |  |  |  |  |  |
| --- | --- | --- | --- | --- | --- | --- | --- | --- | --- | --- | --- | --- | --- | --- | --- | --- |
|  | Sample<br>size | 2.5 | 5 | 25 | 50 | 75 | 95 | 97.5 | Sample<br>size | 2.5 | 5 | 25 | 50 | 75 | 95 | 97.5 |
| 12 | 61 | 3.3 | 3.4 | 3.8 | 4.0 | 4.3 | 4.8 | 5.0 | 65 | 3.1 | 3.2 | 3.5 | 3.8 | 4.0 | 4.5 | 4.7 |
| 13 | 91 | 3.2 | 3.4 | 3.7 | 3.9 | 4.2 | 4.7 | 4.9 | 89 | 3.0 | 3.1 | 3.4 | 3.6 | 3.8 | 4.3 | 4.5 |
| 14 | 87 | 3.2 | 3.3 | 3.6 | 3.8 | 4.1 | 4.6 | 4.8 | 96 | 2.9 | 3.0 | 3.3 | 3.5 | 3.7 | 4.2 | 4.4 |
| 15 | 93 | 3.1 | 3.2 | 3.5 | 3.7 | 4.0 | 4.4 | 4.6 | 76 | 2.8 | 2.9 | 3.2 | 3.4 | 3.6 | 4.1 | 4.3 |
| 16 | 91 | 3.0 | 3.1 | 3.4 | 3.7 | 3.9 | 4.3 | 4.5 | 84 | 2.7 | 2.8 | 3.1 | 3.3 | 3.5 | 4.0 | 4.2 |
| 17 | 91 | 3.0 | 3.1 | 3.4 | 3.6 | 3.8 | 4.2 | 4.4 | 78 | 2.7 | 2.8 | 3.1 | 3.3 | 3.5 | 4.0 | 4.2 |
| 18 | 72 | 2.9 | 3.0 | 3.3 | 3.5 | 3.7 | 4.1 | 4.3 | 69 | 2.8 | 2.9 | 3.2 | 3.4 | 3.6 | 4.1 | 4.3 |

Abbreviations: TSH= thyroid stimulating hormone; T4 = thyrotropin; T3= thyroxine
SI units conversion: Total T4 (nmol/L) = multiply by 12.9; Total T3 (nmol/L) = multiply by 0.0154; fT4 (pmol/L) = multiply by 12.9; fT3 (pmol/L) = multiply by 1.5362

## Discussion

This is the first study to comprehensively examine the association of TSH levels with CMRF in children from a nationally representative non-institutionalized population in the United States comprising of 5 NHANES cycles over 10 years. The prevalence of SH was low (2.0%, 95% CI 1.2-3.1) in this study with no differences in children with normal BMI or those with overweight and obesity. This is remarkably different from the report of SH in 14.2% children in the Korean National Health and Nutrition Examination Surveys reported by Lee *et al* with statistically significant higher proportion in children with obesity (47). Asians are known to have higher cardiometabolic risk compared to other ethnicities at comparable age and sex-adjusted BMI (48-51) and the higher prevalence of SH may reflect similar racial/ethnic differences. Further, it is possible that the higher level of SH may be associated with the presence of anti-thyroid antibodies as no assays were reported for these in their study.

SH is frequently reported in children with overweight and obesity (12,17,18) and one study has shown its resolution with treatment achieved by medical or surgical means (52). The elevated leptin levels seen in obesity and the homeostatic mechanism for preserving body weight are thought to be the mechanisms underlying the elevated TSH levels seen with obesity (53-55). In this study, we showed an association of higher TSH levels with abdominal obesity and insulin resistance after controlling for covariates such as age, gender, race/ethnicity and level of obesity. Animal and human studies have identified a complex relationship of thyrotropin, thyroid hormones and glucose metabolism (56). Whether such relationships are mediated by the direct effects of TSH and thyroid hormones on skeletal muscle and adipose tissue or via hepatic metabolism or mitochondrial oxidation remains to be identified (56,57). Euglycemic-hyperinsulinemic clamp studies in humans have demonstrated decreased insulin-mediated glucose disposal in individuals with hypothyroidism (58) that reverted with treatment (58,59). Whether the reduced peripheral insulin sensitivity in hypothyroid state is caused by the reduction of blood flow to the skeletal muscle and adipose tissue as seen in a study that measured the differences in the arteriovenous sampling in hypothyroid individuals compared to normal controls (60) or other mechanisms such as an increase in glucose uptake by skeletal muscle and adipose tissue demonstrated in an individual with defect in insulin receptor treated with liothyronine (61) remains to be elucidated. In a longitudinal study of health check-up program, Jun *et al* have shown that the risk of incident Type 2 diabetes was significantly increased with each 1 mIU/mL increment in TSH after adjusting for confounding factors and that individuals in the highest tertile of TSH change were at greater risk (62,63). The findings from our study suggest that the association between the higher TSH levels and insulin resistance begins in early adolescence and may need long-term monitoring.

Circulating TSH is an important homeostatic hormone required for maintaining the body milieu and linear association of risk factors with TSH levels is unlikely. The association of cardiometabolic risks with the higher quantile may suggest that perhaps, the currently accepted upper range of normal levels of TSH requires modification. This issue has previously been debated in adult population with the report of mean TSH of 1.50 mIU/L (95% CI 1.46-1.54) in 13,344 people evaluated in NHANES III (64). In a subsequent study, it was reported that the prevalence of TPO antibody was higher in the individuals with TSH levels > 2 mIU/L, especially in those of older age. Therefore, the tail of the distribution with the higher TSH levels may represent occult thyroid dysfunction and an accurate population reference range remains an open question (65,66). The American Thyroid Association currently maintains that in children levothyroxine replacement therapy may be initiated with TSH > 10 mIU/L with signs/symptoms consistent with thyroid disease. It is also noted that if treatment is initiated, the goal of therapy is to maintain the TSH levels in the lower half of reference range, optimally between 0.5-2.0 mIU/L (67). These guidelines leave the range between 2-10 mIU/L without specific guidance, frequently resulting in referral to pediatric endocrinology providers, with disproportionately higher referrals of children with obesity. The debate on whether or not children with SH with or without obesity should be treated continues (68). Whether the higher odds of cardiometabolic risk in the 4th quantile translates into therapeutic relevance can only be determined by longitudinal studies in future.

To provide guidance to the clinicians caring for children, we have calculated the percentiles for the thyroid hormones in youth between 12-18 years of age with BMI between 5-85^th^ percentiles without TPO or TG antibody at the time of assessment. The TSH levels decreased with increasing age from 12 to 18 years, while the Total T4 levels increased and the free T4 levels remained stable across the ages. In a study of Danish children, similar trends were seen, although the absolute levels were higher perhaps reflecting differences in assay or differences between populations (69). The 4^th^ quantile of TSH most closely aligns with the 75th percentile of TSH. Future studies should address the optimal cut-off for TSH levels.

There are several limitations of this study. First, we have combined the results from assays used to assess thyroid function parameters over 5 NHANES cycles over 10 non-contiguous years. While the distribution of the measured hormonal levels overlaps in these 5 cycles, the possibility of differences across platforms remains. The NHANES survey does not include assessment for puberty. We used quantiles adjusted for age and sex to account for this lack of information, that may or may not capture the influence of puberty on thyroid function. We reconstructed the weights from the 5 different NHANES cycles in this analysis. While this estimation is according to the prevalent guidelines from CDC, it is possible that these reconstructed weights may not apply to the population at large at a given time. The cardiometabolic risk factors were ascertained at the same time as the measurement of TSH, hence no inference can be drawn about causation. Finally, the study cohort was ascertained nearly a decade ago. Although we observed no temporal differences in the measurements from 1999-2012, it is possible that the more recent trends may be different.

Notwithstanding these limitations, we believe that this study provides an important reference for clinicians in practice, both for the reference ranges provided as well as the association of CMRF with the highest quantile of TSH levels. Future longitudinal studies are needed to estimate if these CMRFs translate into adverse cardiometabolic outcomes and whether intervention with thyroid replacement therapy to maintain the TSH levels within a certain percentile is warranted.

## Data Availability

The data are publicly available and appropriate references are included in the manuscript.

## Acknowledgements

The authors gratefully acknowledge the critical review of the manuscript by Bat-Sheva Levine, MD, MPH from Boston Children’s Hospital and Stavroula Osganian, MD, ScD from National Institutes of Health (NIH). We are grateful to Qixuan Chen, PhD, from the Mailman School of Public Health at Columbia University for biostatistical consultation. We thank Morten Asp Vonsild Lund, MD PhD from the Department of Biomedical Sciences at University of Copenhagen for sharing their R code. This publication was supported in part by the National Institute for Diabetes and Digestive and Kidney Diseases, NIH, through Grant Number K23 DK110539 and the National Center for Advancing Translational Sciences, NIH, through Grant Number UL1TR001873. The content is solely the responsibility of the authors and does not necessarily represent the official views of the NIH.

